# Effects of intervention stage completion in an integrated behavioral health and primary care randomized pragmatic intervention trial

**DOI:** 10.1101/2024.02.07.24302481

**Authors:** Kari A. Stephens, Constance van Eeghen, Zihan Zheng, Tracy Anastas, Kris Pui Kwan Ma, Maria G. Prado, Jessica Clifton, Gail Rose, Daniel Mullin, Kwun C. G. Chan, Rodger Kessler

**Author notes:** No co-authors have a conflict of interest.

## Abstract

**Purpose:** A pragmatic, cluster-randomized controlled trial of a comprehensive practice-level, multi-staged practice transformation intervention aimed to increase behavioral health integration in primary care practices and improve patient outcomes. We examined association between the completion of intervention stages and patient outcomes across a heterogenous national sample of primary care practices.

**Methods:** Forty-two primary care practices across the U.S. with co-located behavioral health and 2,426 patients with multiple chronic medical and behavioral health conditions completed surveys at baseline, midpoint and two year follow-up. Effects of the intervention on patient health and primary care integration outcomes were examined using multilevel mixed-effects models, while controlling for baseline outcome measurements.

**Results:** No differences were found associated with the number of intervention stages completed in patient health outcomes were found for depression, anxiety, fatigue, sleep disturbance, pain, pain interference, social function, patient satisfaction with care or medication adherence. The completion of each intervention stage was associated with increases in Practice Integration Profile (PIP) domain scores and were confirmed with modeling using multiple imputation for: Workflow 3.5 (95% CI: 0.9-6.1), Integration Methods 4.6 (95% CI: 1.5-7.6), Patient Identification 2.9 (95% CI: 0.9-5.0), and Total Integration 2.7 (95% CI: 0.7-4.7).

**Conclusion:** A practice-centric flexible practice transformation intervention improved integration of behavioral health in primary care across heterogenous primary care practices treating patients with multiple chronic conditions. Interventions that allow practices to flexibly improve care have potential to help complex patient populations. Future research is needed to determine how to best target patient health outcomes at a population level.

## INTRODUCTION

Most patients with behavioral health problems only receive behavioral care in primary care settings rather than specialty mental health settings^1^ and 40% of patients seen in primary care settings have behavioral health needs^2^ with an estimated 30%-80% of primary care visits relating to behavioral health issues.^3^ Primary care struggles to address these complex needs with only 26%-44% of primary care practices having a co-located behavioral health provider (BHP),^4^ although these numbers are increasing.^5^

Integrated Behavioral Health (IBH) is associated with improved access and engagement in mental health services, mental and physical health patient outcomes, and experience of care.^6–10^ IBH models vary^6,7^ and typically they include having a BHP, such as a psychologist or social worker, embedded into the primary care practice, who work collaboratively with primary care providers to assess and manage behavioral health needs.^11^

Evidence-based IBH models of care are difficult to implement, given they require complex practice-level changes customized to each practice and interventions and impact vary.^6,7,12–14^ Best practices for exemplary integration have been identified that include a clear mission and focus on behavioral health, quality improvement processes, defining clear staff and clinician roles, and a team-based approach.^15^ Practice facilitation, Lean Management approaches, and learning collaboratives have specifically been shown to facilitate IBH implementation.^16–18^

The current study tested a pragmatic, cluster-randomized controlled trial to evaluate a comprehensive practice-level intervention to improve behavioral health integration and patient outcomes in primary care practices, specifically targeting patients with multiple (two or more) chronic medical and behavioral health conditions. Primary care practices across the U.S. were randomized to either a control arm of treatment as usual versus a 24-month intervention arm that tested a multi-staged practice-based intervention informed by a Lean Management Toolkit, a structured redesign method to improve IBH with optional process improvement workbooks, quality improvement coaching, clinician and staff education, and collaborative learning. We hypothesized that practices that completed more stages in the intervention arm would report higher levels of integration and patients in these practices would report greater improvement in their physical and mental health over time.

## METHODS

### Study Design

Primary care practices were randomized into one of two arms within a large-scale, pragmatic, cluster-randomized, clinical trial to test the intervention. The active intervention arm, which included a toolkit-based implementation strategy to increase the degree of IBH, was compared to the treatment as usual arm. The study protocol is further detailed elsewhere,^19^ registered at ClinicalTrials.gov NCT02868983 and approved by the University of Vermont Committees on Human Subjects (CHRMS #16- 554) and Institutional Review Boards at other participating locations.

### Practices and Participants

Eligible primary care practices were required to have an existing employed and co-located BHP of 0.5 full-time equivalent (FTE) or more, actively bill Medicare and other insurers for BHP services, use a shared electronic health record system, and score below 75 out of 100 on the Practice Integration Profile (PIP)^20^ to ensure room for improvement in integration was warranted. Eligible patients had at least one chronic medical condition and at least one chronic behavioral health condition or three or more medical conditions.

### IBH and Primary Care (IBH-PC) Intervention

Practices randomized to the active intervention group were provided with the IBH-PC Toolkit, which included four components: 1) workbooks to guide the quality improvement (QI) project; 2) online education tailored to practice personnel roles (medical provider, BHP, nurse, *etc.*); 3) an online learning community; and 4) remote coaching for the primary care practice’s QI team facilitator and QI team by a trained quality improvement (QI) professional paired with a psychologist familiar with IBH. Portions of the Toolkit were iteratively developed in previous studies.^16,21–23^ In keeping with the pragmatic design of this study, each team tailored its use of the Toolkit according to the needs of the practice, including determining when to start within a 2-year timeframe and which components of the intervention to use.

The Toolkit was presented in stages to organize the QI team’s activities into discrete steps to move towards a higher degree of IBH: Stage 1 – planning; Stage 2 – redesign of workflows, and Stage 3 – implementation of practice changes. The education, online learning, and remote coaching components were offered throughout the three stages and were accessed as needed by each primary care practice-based QI team. Each stage included a set of steps for QI teams to follow and coaches confirmed progress across steps and assisted in adaptations to the intervention to best meet the team’s goals as teams requested coaching support. Coaches documented completion of each step or completion of an adapted step. For example, an early step in Stage 1 (Planning) was “Develop your vision of IBH,” where an intervention arm practice chose to review and revise a recently developed vision statement regarding IBH, after which the coach documented that step as “complete.” Coaches met weekly as a coaching group to review practice progress and come to consensus on coding of completion of steps, which were documented in a shared record. Practices that had completed all steps in a stage were considered to have completed that stage for analysis purposes. Practices that chose to skip one or more steps were re- assessed by coaches separately, based on coaching notes, and then cross-compared and finalized in follow-up coaching team meetings to reach final consensus on stage completion status. If one or more steps in a stage was assessed as “skipped,” that stage was deemed to be incomplete.

### Measures

All measures were administered by surveys to practice and patient participants at baseline, midpoint, and the 2-year timepoints.^19^ Participants provided baseline data directly after practice randomization or patient recruitment, midpoint data at approximately 12-18 months after baseline, and 2-year data at approximately 21-27 months after baseline.

#### Patient Heath Outcomes (PROMIS-29, PHQ-9, GAD-7)

The PROMIS-29 measured patients’ physical function, anxiety, depression, fatigue, sleep disturbance, social participation, and pain interference in the past seven days, using a 5-point response option with each separate scale.^24,25^ An additional pain numerical rating scale (0-10) was included where a higher rating indicated more intense pain. PROMIS-29 items were also used to create composite scores of mental and physical health summary scores.^26^ Responses were scored on a T-score metric based on the PROMIS normative reference sample of U.S. adults, scales scored with a mean equal to 50 and a standard deviation of 10. A higher T-score indicated worse severity in anxiety, depression, fatigue, pain interference, pain intensity, and sleep disturbance. A lower T-score indicated worse severity in physical function and social participation. Scores 3 or more away from 50 indicated at least mild impairment. Depression was measured by the PHQ-9, a self-administered, screening tool for assessment of the severity of depressive symptoms.^27^ The PHQ-9 has good reliability (*α* = 0.89). (Anxiety was measured by the GAD-7, a brief 7- item self-report scale that identifies probable cases of general anxiety disorder (GAD).^28^ The GAD-7 has good reliability (*α* = 0.83).

#### Practice Integration Outcomes

The Practice Integration Profile (PIP) version 1.0 was developed by a national team of clinicians and clinical researchers and operationalizes the Lexicon of Integrated Care.^29^ The PIP has been shown to discriminate differing levels of integrated care processes and differences in type of practice.^20^ The 30-item PIP was administered to at least four people at the practice (i.e., a medical primary care provider (PCP), BHP, an administrator such as a clinic manager, and a provider or staff of the practice’s choice). The PIP assessed levels of the practice’s behavioral health integration across six domains: practice workflow, clinical services, integration methods, case identification, patient engagement, and workspace arrangement and infrastructure. For each domain, the scores range from 0 (no integration) to 100 (full integration). The Total Integration PIP Score is the unweighted average of the six-domain scores.^20,30,31^

### Statistical Analysis

Multilevel mixed-effects models were conducted using the number of intervention stages completed as the primary exposure of interest. Baseline outcome measurement, as well as the time interval from baseline to midpoint and follow-up measurements, respectively, were adjusted for in all analyses.

#### Patient Health Outcomes

We evaluated the association between the number of IBH-PC intervention stages completed and patient reported outcomes using 3-level mixed models with repeated (midpoint and 2-year follow-up) measurements (level 1) nested in patients (level 2) nested in individual primary care practices (level 3). Patient and practice were modeled as random effects. Each model included two random intercepts to account for the difference in average outcome at individual and practice levels. For categorical outcomes, only patients with room for improvement were analyzed (i.e., baseline measures for PROMIS-29 were >=2 points worse than the 50, PHQ-9 >= 10, GAD>=10; and patients reported associated condition, e.g., depression, anxiety, etc.). We adjusted for age, sex, race, ethnicity, employment status, living region (urban/rural), and insecurity status (i.e., noted as present if at least one food, housing, or financial deprivation was reported). Socioeconomic disadvantage was included as a binary variable (i.e., present, not present) if either food, housing or finance was insecure.

#### Practice Integration Outcomes

We assessed the association between the number of IBH-PC intervention stages completed and PIP total and scale scores using 3-level mixed models with repeated (midpoint and 2-year follow-up) measurements (level 1) nested in staff/providers (level 2) nested in primary care practices (level 3). Staff/provider and practice were modeled as random effects. Each model included two random intercepts to account for the difference in average PIP score at individual and practice levels. We adjusted for the ratio of BHP FTE: PCP FTE, baseline outcome measurement, as well as the time interval from baseline to midpoint and follow-up measurements, respectively.

#### Sensitivity Analysis

To test the robustness of the results, we performed multiple imputations by chained equations using the mice function in R,^32^ in addition to complete-case analysis, to handle the missing data across three time points caused by participant non-response. We used 25 imputations and 30 iterations to predict missing data values. The intervention effects on patient-reported outcomes and practice integration were assessed using the methods described above with exclusion of one practice that did not complete any intervention stages.

## RESULTS

A total of 42 practices were randomized in the study with one practice unable to provide eligible patient data and therefore not included in the patient level analyses (see Figure 1). All primary care providers and staff (*N* = 237) and patients (*N* = 2,945) completed baseline and at least one of the follow-up assessments. Patients were on average older (*M* age = 61.9), majority female (65.5%), with an average of 4.4 chronic conditions at baseline, and some patient baseline characteristics related to race, annual household income, diabetes, and urban/rural area significantly differed between study arms (see Table 1). The primary care practices were predominantly non-profit organizations (88%), located in urban areas (83%) and had no significant differences in practices characteristics between arms. Among the 20 primary care practices randomized to the intervention arm, 13 (65%) practices completed all three intervention stages, 6 (30%) practices completed two stages, and 1 (5%) practice did not complete any stage.

**Figure 1.**
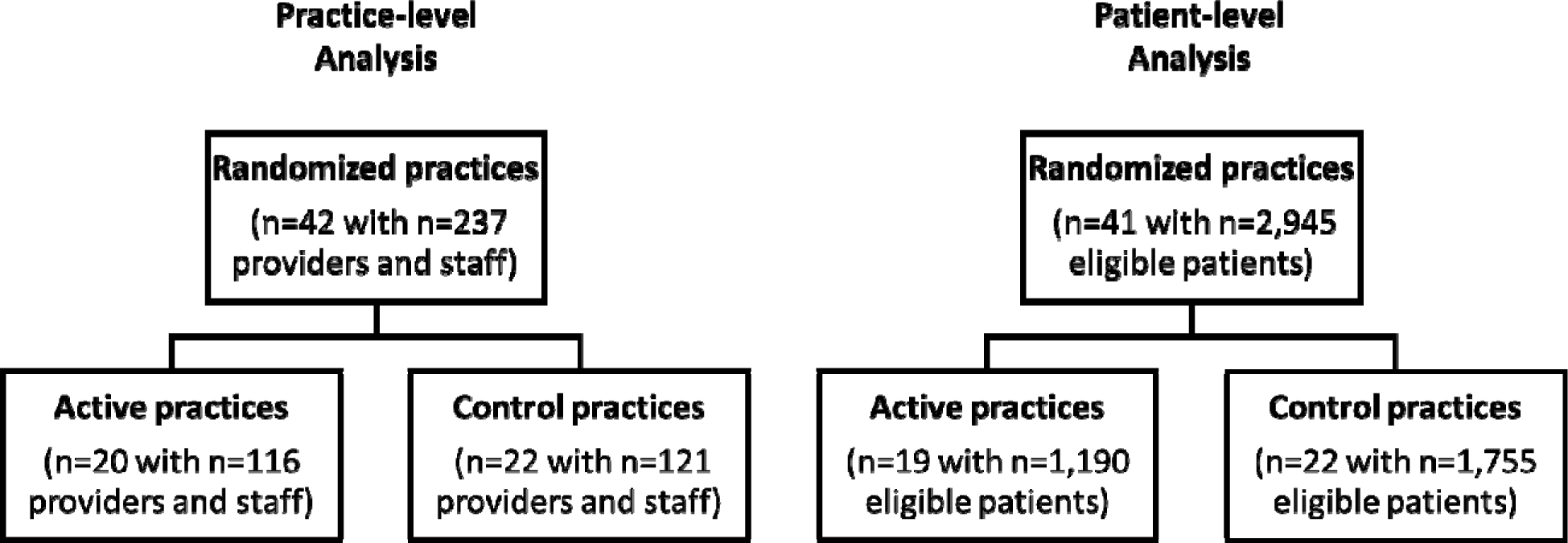
CONSORT Diagram.

**Table 1.** Patient characteristics and outcomes at baseline.

|  | Overall<br>(N=2945) | Active Site<br>(N=1190) | Control Site<br>(N=1755) | p-value |
| --- | --- | --- | --- | --- |
| **Age, years** | 61.8 (13.3) | 61.7 (12.8) | 61.8 (13.6) | 0.90 |
| **Sex** |  |  |  | 0.11 |
| Female | 1884 (64.0%) | 742 (62.4%) | 1142 (65.1%) |  |
| Male | 1054 (35.8%) | 448 (37.6%) | 606 (34.5%) |  |
| **Race** |  |  |  | 0.001* |
| White | 2215 (75.2%) | 849 (71.3%) | 1366 (77.8%) |  |
| Black or African American | 356 (12.1%) | 160 (13.4%) | 196 (11.2%) |  |
| American Indian or Alaskan Native | 30 (1.0%) | 12 (1.0%) | 18 (1.0%) |  |
| Asian | 94 (3.2%) | 51 (4.3%) | 43 (2.5%) |  |
| Native Hawaiian/Other Pacific Islander | 42 (1.4%) | 23 (1.9%) | 19 (1.1%) |  |
| Other/Prefer not to say | 202 (6.9%) | 91 (7.6%) | 111 (6.3%) |  |
| **Ethnicity** |  |  |  | 0.70 |
| Hispanic | 240 (8.1%) | 103 (8.7%) | 137 (7.8%) |  |
| Non-Hispanic | 2660 (90.3%) | 1068 (89.7%) | 1592 (90.7%) |  |
| Prefer not to say | 29 (1.0%) | 12 (1.0%) | 17(1.0%) |  |
| **Marital Status** |  |  |  | 0.06 |
| Never married | 489 (16.6%) | 186 (15.6%) | 303 (17.3%) |  |
| Married | 1286 (43.7%) | 550 (46.2%) | 736 (41.9%) |  |
| Living as married | 76 (2.6%) | 26 (2.2%) | 50 (2.8%) |  |
| Separated | 81 (2.8%) | 23 (1.9%) | 58 (3.3%) |  |
| Divorced | 638 (21.7%) | 260 (21.8%) | 378 (21.5%) |  |
| Widowed | 366 (12.4%) | 141 (11.8%) | 225 (12.8%) |  |
| **Employment** |  |  |  | 0.02* |
| Full-time | 562 (19.1%) | 249 (20.9%) | 313 (17.8%) |  |
| Part-time | 240 (8.1%) | 85 (7.1%) | 155 (8.8%) |  |
| Retired | 1029 (34.9%) | 407 (34.2%) | 622 (35.4%) |  |
| Disabled | 783 (26.6%) | 316 (26.6%) | 467 (26.6%) |  |
| Homemaker | 107 (3.6%) | 39 (3.3%) | 68 (3.9%) |  |
| Student | 20 (0.7%) | 2 (0.2%) | 18 (1.0%) |  |
| Unemployed/Looking for jobs | 94 (3.2%) | 33 (2.8%) | 61 (3.5%) |  |
| Other | 4 (0.1%) | 1 (0.1%) | 3 (0.2%) |  |
| **Annual household income** |  |  |  | 0.02* |
| <\$15,000 | 871 (29.6%) | 344 (28.9%) | 527 (20.0%) | |
| \$15,000-\$29,999 | 625 (21.2%) | 249 (20.9%) | 376 (30.0%) | |
| \$30,000-\$44,999 | 333 (11.3%) | 121 (10.2%) | 212 (12.1%) | |
| \$45,000-\$59,999 | 215 (7.3%) | 81 (6.8%) | 134 (7.6%) | |
| \$60,000-\$74,999 | 219 (7.4%) | 85 (7.1%) | 134 (7.6%) | |
| \$75,000+ | 533 (18.1%) | 253 (21.3%) | 280 (16.0%) | |
| **Education** |  |  |  | 0.98 |
| Less than 9th grade | 84 (2.9%) | 36 (3.0%) | 48 (2.7%) |  |
| 9th to 12th grade, no diploma | 271 (9.2%) | 110 (9.2%) | 161 (9.2%) |  |
| High school graduate (including GED) | 1217 (41.3%) | 485 (40.8%) | 732 (41.7%) |  |
| Associate degree | 450 (15.3%) | 185 (15.5%) | 265 (15.1%) |  |
| Bachelor's degree | 446 (15.1%) | 186 (15.6%) | 260 (14.8%) |  |
| Graduate or professional degree | 406 (13.8%) | 162 (13.6%) | 244 (13.9%) |  |
| **Chronic Conditions** |  |  |  |  |
| Arthritis | 1239 (42.1%) | 505 (42.4%) | 734 (41.8%) | 0.77 |
| Asthma | 650 (22.1%) | 276 (23.2%) | 374 (21.3%) | 0.24 |
| Chronic Obstructive Pulmonary Disease | 422 (14.3%) | 162 (13.6%) | 260 (14.8%) | 0.36 |
| Chronic pain | 2037 (84.0%) | 824 (85.2%) | 1213 (83.1%) | 0.17 |
| Non-Gestational Diabetes | 1335 (45.3%) | 512 (43.0%) | 823 (46.9%) | 0.04* |
| Heart failure | 243 (8.3%) | 96 (8.1%) | 147 (8.4%) | 0.80 |
| Hypertension | 2434 (82.6%) | 999 (83.9%) | 1435 (81.8%) | 0.13 |
| Irritable bowel syndrome | 127 (4.3%) | 61 (5.1%) | 66 (3.8%) | 0.08 |
| Anxiety | 1016 (34.5%) | 420 (35.3%) | 596 (34.0%) | 0.48 |
| Depression | 1418 (48.1%) | 570 (47.9%) | 848 (48.3%) | 0.85 |
| Insomnia | 734 (24.9%) | 314 (26.4%) | 420 (23.9%) | 0.14 |
| Substance use disorder | 714 (24.2%) | 299 (25.1%) | 415 (23.6%) | 0.38 |
| Tobacco use | 574 (19.5%) | 233 (19.6%) | 341 (19.4%) | 0.92 |
| Alcohol use disorder | 201 (6.8%) | 88 (7.4%) | 113 (6.4%) | 0.32 |
| Mean number of chronic conditions | 4.4 (1.7) | 4.5 (1.7) | 4.4 (1.6) | 0.17 |
| **Neighborhood characteristics (home census tract)** |  |  |  |  |
| Social Deprivation Index | 53.5 (27.8%) | 52.4 (29.1%) | 54.2 (27.0%) | 0.09 |
| Urban | 2329 (79.1%) | 989 (83.1%) | 1340 (76.4%) | <0.001* |
| Population density, persons per square mile | 3900 (6670) | 5130 (9510) | 3090 (3350) | <0.001* |
| **Food insecurity** | 366 (12.4%) | 130 (10.9%) | 236 (13.4%) | 0.04* |
| **Housing insecurity** | 97 (3.3%) | 37 (3.3%) | 60 (3.4%) | 0.68 |
| **Financial insecurity** | 697 (23.7%) | 266 (22.4%) | 431 (24.6%) | 0.14 |
| **Drinking category** |  |  |  | 0.78 |
| Non-drinker | 1264 (42.9%) | 466 (39.2%) | 798 (45.5%) |  |
| Drinker | 673 (22.9%) | 240 (20.2%) | 433 (24.7%) |  |
| Unsafe drinker | 205 (7.0%) | 78 (6.6%) | 127 (7.2%) |  |
| **Primary Outcomes –**<br>**PROMIS-29 t-scores** |  |  |  |  |
| Anxiety | 54.1 (10.1) | 54.2 (10.1) | 54.0 (10.1) | 0.77 |
| Depression | 53.0 (9.8) | 52.9 (9.7) | 53.0 (10.0) | 0.85 |
| Fatigue | 52.7 (10.4) | 52.5 (10.3) | 52.8 (10.4) | 0.42 |
| Sleep Disturbance | 53.2 (8.9) | 53.3 (9.0) | 53.2 (8.9) | 0.64 |
| Pain Interference | 58.3 (10.1) | 58.3 (10.0) | 58.4 (10.2) | 0.77 |
| Pain Intensity | 4.5 (2.8) | 4.5 (2.8) | 4.5 (2.8) | 0.98 |
| Social Participation* | 48.1 (10.0) | 48.1 (10.0) | 48.1 (10.1) | 0.85 |
| Physical Function* | 43.2 (9.5) | 43.5 (9.4) | 43.0 (9.5) | 0.17 |
| **Secondary Outcomes** |  |  |  |  |
| PROMIS-29 Physical<br>Health<br>Summary t-score* | 45.5 (9.5) | 45.8 (9.4) | 45.3 (9.6) | 0.22 |
| PROMIS-29 Mental<br>Health<br>Summary t-score* | 50.1 (8.9) | 50.2 (8.9) | 50.1 (8.9) | 0.76 |
| CARE total score* | 4.3 (0.9) | 4.3 (0.9) | 4.2 (0.9) | 0.09 |
| MMAS total score* | 1.0 (1.2) | 1.1 (1.2) | 1.0 (1.1) | 0.46 |
| Metabolic Equivalents | 6.4 (2.0) | 6.4 (2.0) | 6.4 (2.0) | 0.87 |
| PHQ-9 total score | 6.6 (6.1) | 6.6 (6.2) | 6.5 (6.1) | 0.84 |
| GAD-7 total score | 4.7 (5.3) | 4.8 (5.4) | 4.6 (5.3) | 0.38 |
| Asthma Symptom Utility<br>Index | 0.8 (0.2) | 0.8 (0.2) | 0.8 (0.2) | 0.74 |

Note: □ Higher score is better; \* $p < .05$ ; $p$ -value for two-sample t-test used for continuous variables and Chi-squared tests used for categorical variables.

### Patient Health Outcomes

Patients with multiple chronic conditions in the intervention arm did not report significantly different outcomes compared to patients in the treatment as usual arm. Namely, no significant association was found between the number of intervention stages completed and patient health outcomes (see Figure 2), consistent with the sensitivity analysis. These analyses did not account for amount of service accessed by medical or behavioral health providers at each practice.

**Figure 2.**
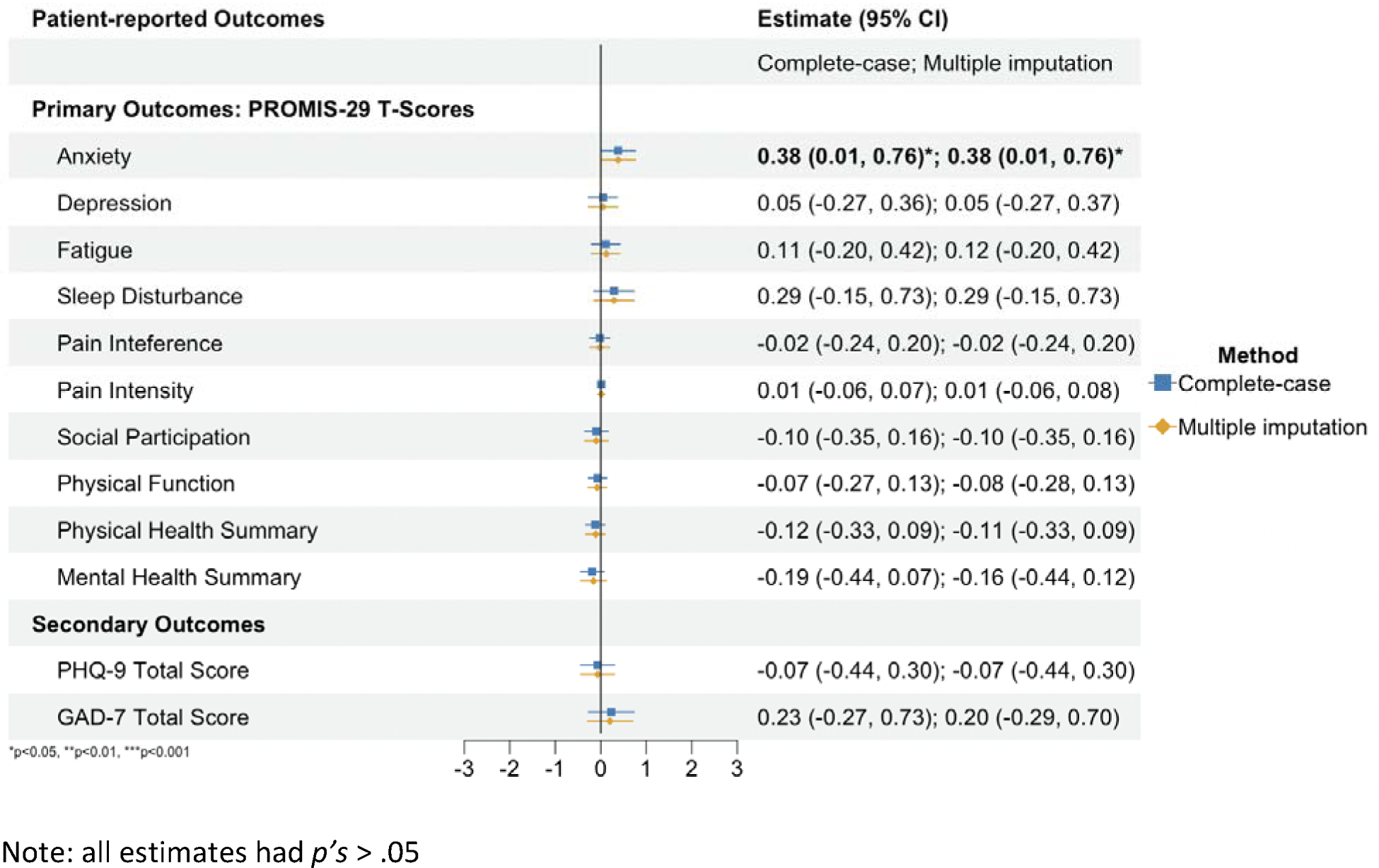
Adjusted effect of intervention stage completion on patient health outcomes.

### Practice Integration Outcomes

Primary care practice personnel in the intervention arm reported significantly higher integration based on the PIP compared to the practices in the treatment as usual arm. With complete-case analysis, the completion of each intervention stage was associated with a significant increase in PIP scores of 3.5 (95% CI: 0.9-6.1) for Workflow, 4.6 (95% CI: 1.5-7.6) for Integration Methods, 2.9 (95% CI: 0.9-5.0) for Patient Identification, and 2.7 (95% CI: 0.7-4.7) for Total Integration PIP score (see Figure 3). After implementing multiple imputation, results patterns remained consistent (see Figure 3).

**Figure 3.**
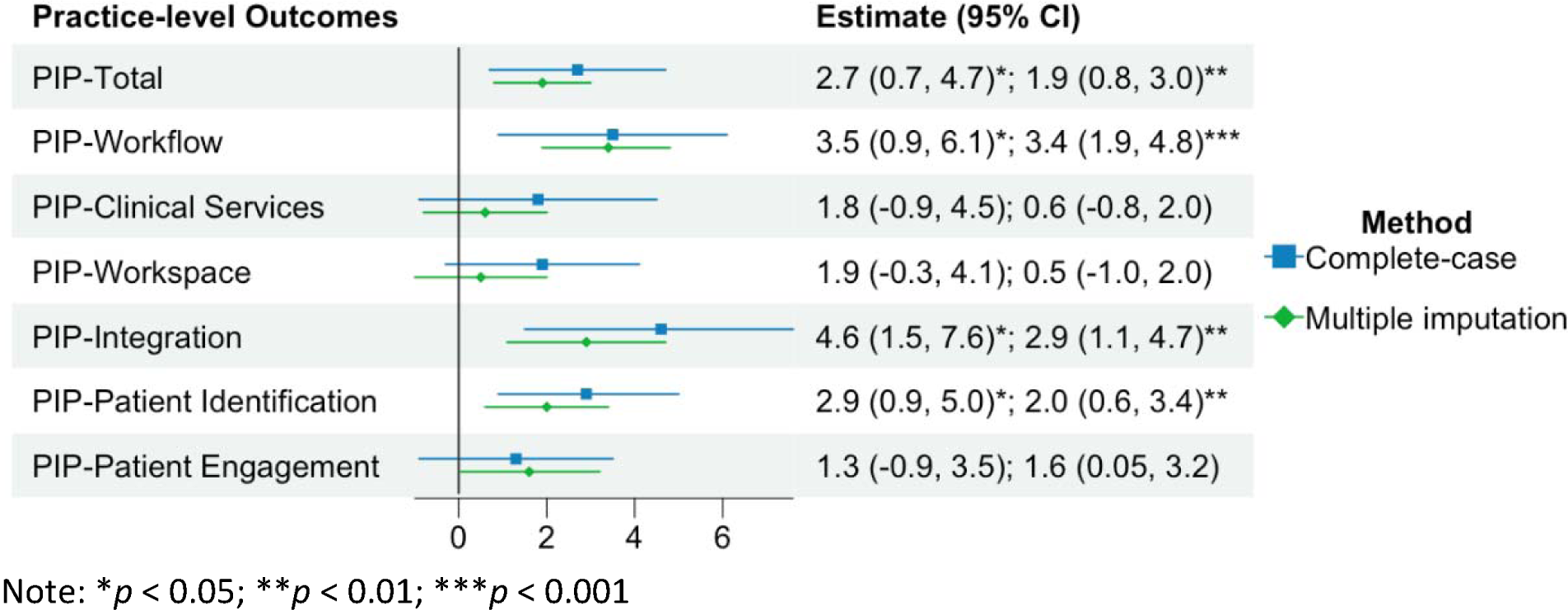
Adjusted effect of intervention stage completion on integration level.

## DISCUSSION

A QI toolkit designed to allow practices to define practice-centric targets for improvement, offered an effective method for busy, complex primary care practices to significantly increase overall level of integration, as well as across workflow, integration methods, and patient identification. Although Improvements in the practice of integration were likely modest, given the increases in integration scores were small, practice level changes were complex, practice-driven and some changes occurred during the disruptions of the 2020 COVID pandemic. Practice-centric approaches to implementing IBH help practices, given modest changes to practice require a substantial investment in time and resources and are necessary for implementation of substantial and sustainable change.

Although patient health outcomes across a random sample of patients with multiple chronic medical and behavioral health conditions were not observed, analyses did not select or measure engagement in direct care by BHPs (i.e., patients may not have received any direct treatment from the BHP) and patients may have already received treatment for their chronic conditions at time of baseline measurement. Future studies are needed to evaluate whether patient outcomes may improve who have direct engagement in care with BHPs, even with modest improvements in IBH.

The success of the intervention was likely due to the practice-centric approach that allowed practices to flexibly set custom goals to their settings, using evidence-based QI methods and targeting a defined group of patients with multiple chronic medical and behavioral health conditions. Practices were able to designate their own intervention teams, meet on their own choice of schedule and frequency and engage materials and resources as they saw fit to make real change in their practices of integration. Given the variation in primary care practices’ readiness and capacity for change, as well as the heterogeneity inherent across these practices’ processes and structures, practice-centric interventions that balance flexibility with consistent structure such as this intervention may help reduce barriers to disseminating IBH. The demand to meet the needs of the mental health wave from the COVID-19 pandemic and the continued rise in chronic diseases, which are the leading cause of death and disability in the U.S. can often be prevented and treated through evidence-based behavioral interventions.^33^

Changes in integration did not affect clinical services, workspace, or patient engagement domains. Changes in clinical services and patient engagement may be more difficulty to measure with a quick self report measure, as well as be more difficult to effect in a practice if specific goals related set by the practices did not target these areas for improvement. Workspace was rated as the highest domain of integration (see Table 2) and was high (87.6 out of 100) at baseline and therefore had less room for improvement.

**Table 2.** Practice characteristics and outcomes at baseline.

|  | Overall<br>(N = 42) | Intervention Arm<br>(n = 20) | Control Arm<br>(n = 22) | *p-value* |
| --- | --- | --- | --- | --- |
| **Practice specialty** |  |  |  | 0.85 |
| Internal medicine | 7 (17%) | 3 (15%) | 4 (18%) |  |
| Family medicine | 20 (48%) | 9 (45%) | 11 (50%) |  |
| Mixed | 15 (36%) | 8 (40%) | 7 (32%) |  |
| **Organization type** |  |  |  |  |
| Community Health Center | 15 (36%) | 8 (40%) | 7 (32%) | 0.82 |
| Hospital | 20 (48%) | 10 (50%) | 10 (45%) | 0.77 |
| Private | 4 (10%) | 1 (5%) | 3 (14%) | 0.67 |
| Academic | 19 (45%) | 10 (50%) | 9 (41%) | 0.78 |
| **Resident training site** | 16 (38%) | 9 (45%) | 7 (32%) | 0.58 |
| **Non-profit** | 37 (88%) | 19 (95%) | 18 (82%) | 0.40 |
| **Geographic region** |  |  |  | 0.88 |
| Pacific Northwest | 3 (7%) | 1 (5%) | 2 (9%) |  |
| Mountain | 8 (19%) | 4 (20%) | 4 (18%) |  |
| The South | 8 (19%) | 4 (20%) | 4 (18%) |  |
| New England | 9 (21%) | 3 (15%) | 6 (27%) |  |
| Mid-Atlantic & Great Lakes | 6 (14%) | 3 (15%) | 3 (14%) |  |
| West Coast & Hawaii | 8 (19%) | 5 (25%) | 3 (14%) |  |
| **Urban by RUCA** | 35 (83%) | 18 (90%) | 17 (77%) | 0.49 |
| **County social deprivation index** | 44.9 (22.0) | 46.4 (23.3) | 43.5 (21.1) | 0.68 |
| **Patient cared for by the practice each year** | 9285 (5066) | 9138 (4549) | 9419 (5599) | 0.86 |
| **Baseline BHP FTE** | 1.5 (1.1) | 1.7 (1.4) | 1.3 (0.7) | 0.18 |
| **Baseline PCP FTE** | 6.0 (3.2) | 5.9 (2.7) | 6.1 (3.6) | 0.83 |
| **Baseline BHP FTE: PCP FTE** | 0.30 (0.26) | 0.35 (0.28) | 0.26 (0.24) | 0.27 |
| **Baseline PIP Total** | 61.0 (17.4) | 60.4 (15.6) | 61.7 (19.1) | 0.62 |
| PIP - workflow | 50.3 (21.9) | 48.7 (20.8) | 51.9 (23.0) | 0.35 |
| PIP - clinical services | 60.4 (21.1) | 61.8 (19.8) | 59.0 (22.5) | 0.41 |
| PIP - workspace | 87.6 (18.2) | 87.3 (15.2) | 87.9 (20.9) | 0.82 |
| PIP - integration | 53.6 (23.3) | 52.1 (21.6) | 55.1 (24.9) | 0.42 |
| PIP - patient identification | 66.8 (22.6) | 65.4 (21.4) | 68.1 (23.8) | 0.46 |
| PIP - patient engagement | 47.5 (22.8) | 46.8 (21.1) | 48.2 (24.5) | 0.70 |

Note: \* $p < .05$ ; *p-value* for two-sample t-test used for continuous variables and Chi-squared tests used for categorical variables.

Primary care practices have struggled to adopt IBH for various reasons that include workforce supply issues with a shortage of BHPs available and willing to work in primary care settings, unstable reimbursement and funding models, little adaptation of behavioral health into practice workflow, and the always present difficulty creating and sustaining practice change.^5,34,35^ Policy changes such as the Affordable Care Act have caused increases in volume of care within community health clinics and other primary care settings, and an increased focus on complexity of the behavioral health issues in need of care in these settings. The COVID-19 pandemic has complicated delivery of primary care through social distancing and exacerbated issues with loneliness, anxiety, and insomnia along with mental health acuity.^36–38^ Results of this intervention demonstrate that despite practices flexing how and when they approach and target practice change, they can nimbly advance IBH when given the right support at a time where patients needs are only getting higher.

Limitations of this study include a sample of practices that already had an established history of integrating behavioral and primary care and scored in the middle range of self-reported IBH as assessed by the PIP. While we did not test whether this intervention is effective with lower levels of integration or with practices just beginning to add BHPs to their practice teams, the flexible nature of this intervention may benefit those practices as well. We also did not account for the quality and quantity of medical care access each patient received, whether the participant was treated specifically by the BHP in the practice, or whether the patient was in need of behavioral interventions at baseline. Future studies are needed with careful patient selection to examine impact of improving IBH on these complex patient populations.

Primary care practices face an unprecedented challenge in demand for care to address the needs among patients with multiple chronic conditions that have only grown during the COVID-19 pandemic. A practice-centric flexible intervention aimed at improving the level of IBH in primary care can help practices transform to meet these needs and improve the health of their most complex patients. Future research is needed to determine how to leverage data collected in routine care (i.e., via electronic health record systems) from patients, to enable careful evaluation of the impact of these primary care transformations and ensure pathways to dissemination of IBH are supported.

## Data Availability

All data produced in the present study are available upon reasonable request to the authors

## Acknowledgements

The authors acknowledge the work of the Integrating Behavioral Health and Primary Care (IBH-PC) Research Team in the implementation of this intervention study. This research was supported by the Patient-Centered Outcomes Research Institute (PCS-1409-24372), the Institute of Translational Health Sciences (UL1 TR002319), and the Primary Care Psychiatry/Behavioral Health Integration Fellowship T32 (T32MH020021).

## Abbreviations

ASUI: Asthma Symptom Utility Index
BHP: Behavioral Health Provider
CARE survey: Consultation and Relational Empathy
FTE: Full-time Equivalent
GAD: Generalized Anxiety Disorder
IBH: Integration of Behavioral Health
IBH-PC: IBH Primary Care
MMAS-8: Morisky Medication Adherence Scale
PCP: Primary Care Provider
PIP: Practice Integration Profile
PROMIS-29: Patient-Reported Outcomes Measurement Information System
QI: Quality Improvement

## References

1. Kessler R, Stafford D. Primary care is the de facto mental health system. In: Kessler R, Stafford D, eds. Collaborative medicine case studies. New York: Springer; 2008. p. 9-21. 10.1007/978-0-387-76894-6_2

2. Ansseau M, Dierick M, Buntinkx F, Cnockaert P, De Smedt J, Van Den Haute M, et al. High prevalence of mental disorders in primary care. J Affect Disord. 2004;78(1):49–55. 10.1016/S0165-0327(02)00219-7

3. Wodarski JS. The integrated behavioral health service delivery system model. Soc Work Public Health. 2014;29(4):301–317. 10.1080/19371918.2011.622243

4. Richman EL, Lombardi BM, Zerden LD. Mapping colocation: using national provider identified data to assess primary care and behavioral health colocation. Fam Syst Health. 2020;38(1):16–23. 10.1037/fsh0000465

5. Tong ST, Morgan ZJ, Stephens KA, Bazemore AW, Peterson LE. Characteristics of family physicians practicing collaboratively with behavioral health professionals. Ann Fam Med. 2023;21(2):157–160. 10.1370/afm.2947

6. Possemato K, Johnson EM, Beehler GP, Shepardson RL, King P, Vair CL, et al. Patient outcomes associated with primary care behavioral health services: a systematic review. Gen Hosp Psychiatry. 2018;53:1–11. 10.1016/j.genhosppsych.2018.04.002

7. Thota AB, Sipe TA, Byard GJ, Zometa CS, Hahn RA, McKnight-Eily LR, et al. Collaborative care to improve the management of depressive disorders: a community guide systematic review and meta-analysis. Am J Prev Med. 2012;42(5):525–538. 10.1016/j.amepre.2012.01.019

8. Archer J, Bower P, Gilbody S, Lovell K, Richards D, Gask L, et al. Collaborative care for depression and anxiety problems. Cochrane Database Syst Rev. 2012;10:CD006525. 10.1002/14651858.cd006525.pub2

9. Balasubramanian BA, Cohen DJ, Jetelina KK, Dickinson LM, Davis M, Gunn R, et al. Outcomes of integrated behavioral health with primary care. J Am Board Fam Med. 2017;30(2):130–139. 10.3122/jabfm.2017.02.160234

10. Katon WJ, Lin EHB, Von Korff M, Ciechanowski P, Ludman EJ, Young B, et al. Collaborative care for patients with depression and chronic illnesses. N Engl J Med. 2010;363(27):2611–2620. 10.1056/nejmoa1003955

11. Beil H, Feinberg RK, Patel SV, Romaire MA. Behavioral health integration with primary care: implementation experience and impacts from the state innovation model round 1 states. Milbank Q. 2019;97(2):543–582. 10.1111/1468-0009.12379

12. Robinson PJ, Strosahl KD. Behavioral health consultation and primary care: lessons learned. J Clin Psychol Med Settings. 2009;16(1):58–71. 10.1007/s10880-009-9145-z

13. Katzelnick DJ, Williams MD. Large-scale dissemination of collaborative care and implications for psychiatry. Psychiatr Serv. 2015;66(9):904–906. 10.1176/appi.ps.201400529

14. Working Party Group on Integrated Behavioral Healthcare. Joint principles: integrating behavioral health care into the patient-centered medical home. Fam Syst Health. 2014;32(2):154–156. 10.1037/h0099809

15. Cohen DJ, Davis MM, Hall JD, Gilchrist EC, Miller BF. A guidebook of professional practices for behavioral health and primary care integration: observations from exemplary sites. Rockville (MD): Agency for Healthcare Research and Quality; 2015. https://integrationacademy.ahrq.gov/sites/default/files/2020-06/AHRQ_AcademyGuidebook.pdf

16. van Eeghen C, Littenberg B, Holman MD, Kessler R. Integrating behavioral health in primary care using lean workflow analysis: a case study. J Am Board Fam Med. 2016;29(3):385–393. 10.3122/jabfm.2016.03.150186

17. Okafor M, Ede V, Kinuthia R, Satcher D. Explication of a behavioral health-primary care integration learning collaborative and its quality improvement implications. Community Ment Health J. 2018;54(8):1109–1115. 10.1007/s10597-017-0230-8

18. Roderick SS, Burdette N, Hurwitz D, Yeracaris P. Integrated behavioral health practice facilitation in patient centered medical homes: a promising application. Fam Syst Health. 2017;35(2):227–237. 10.1037/fsh0000273

19. Crocker AM, Kessler R, van Eeghen C, Bonnell LN, Breshears RE, Callas P, et al. Integrating behavioral health and primary care (IBH-PC) to improve patient-centered outcomes in adults with multiple chronic medical and behavioral health conditions: study protocol for a pragmatic cluster-randomized control trial. Trials. 2021;22(1):200. 10.1186/s13063-021-05133-8

20. Kessler RS, Auxier A, Hitt JR, Macchi CR, Mullin D, van Eeghen C, et al. Development and validation of a measure of primary care behavioral health integration. Fam Syst Health. 2016;34(4):342–356. 10.1037/fsh0000227

21. van Eeghen C, Littenberg B, Kessler R. Chronic care coordination by integrating care through a team-based, population-driven approach: a case study. Transl Behav Med. 2018;8(3):468–480. 10.1093/tbm/ibx073

22. van Eeghen C, Edwards M, Libman BS, MacLean CD, Kennedy AG. Order from chaos: an initiative to improve opioid prescribing in rheumatology using lean A3. ACR Open Rheumatol. 2019;1(9)546-551. 10.1002/acr2.11078

23. van Eeghen C, Kennedy AG, Pasanen ME, MacLean CD. A new quality improvement toolkit to improve opioid prescribing in primary care. J Am Board Fam Med. 2020;33(1):17–26. 10.3122/jabfm.2019.01.190238

24. HealthMeasures. Updated April 4, 2021. Accessed October 26, 2023. https://www.healthmeasures.net/images/PROMIS/manuals/PROMIS_Adult_Profile_Scoring_Manual.pdf

25. Cella D, Riley W, Stone A, Rothrock N, Reeve B, Yount S, et al. The patient-reported outcomes measurement information system (PROMIS) developed and tested its first wave of adult self-reported health outcome item banks: 2005-2008. J Clin Epidemiol. 2010;63(11):1179:1194. 10.1016/j.jclinepi.2010.04.011

26. Hays RD, Spritzer KL, Schalet BD, Cella D. PROMIS((R))-29 v2.0 profile physical and mental health summary scores. Qual Life Res. 2018;27(7):1885–1891. 10.1007/s11136-018-1842-3

27. Blackwell TL, McDermott AN. Test review: patient health questionnaire–9 (PHQ-9). Rehabil Couns Bull. 2014;57(4):246–248. 10.1177/0034355213515305

28. Spitzer RL, Kroenke K, Williams JBW, Löwe B. A brief measure for assessing generalized anxiety disorder: the GAD-7. Arch Intern Med. 2006;166(10):1092–1097. 10.1001/archinte.166.10.1092

29. Peek CJ, National Integration Academy Council. Lexicon for behavioral health and primary care integration: concepts and definitions developed by expert consensus. AHRQ Publication No.13- IP001-EF. Rockville (MD): Agency for Healthcare Research and Quality; 2013. https://integrationacademy.ahrq.gov/sites/default/files/2020-06/Lexicon_ExecSummary.pdf

30. Hitt JR, Brennhofer SA, Martin MP, Macchi CR, Mullin D, van Eeghen C. Further experience with the practice integration profile: a measure of behavioral health and primary care integration. J Clin Psychol Med Settings. 2022;29(2):274–284. 10.1007/s10880-021-09806-z

31. Mullin DJ, Hargreaves L, Auxier A, Brennhofer SA, Hitt JR, Kessler RS, et al. Measuring the integration of primary care and behavioral health services. Health Serv Res. 2019;54(2):379–389. 10.1111/1475-6773.13117

32. White IR, Royston P, Wood AM. Multiple imputation using chained equations: issues and guidance for practice. Stat Med. 2011;30(4):377–399. 10.1002/sim.4067

33. Centers for Disease Control and Prevention [Internet]. Atlanta: Centers for Disease Control and Prevention [cited 2023 Sep 18]. National Center for Chronic Disease Prevention and Health Promotion (NCCDPHP); [about 2 screens]. Available from: https://www.cdc.gov/chronicdisease/index.htm

34. Ma KPK, Mollis BL, Rolfes J, Au M, Crocker A, Scholle SH, et al. Payment strategies for behavioral health integration in hospital-affiliated and non-hospital-affiliated primary care practices [published correction appears in Transl Behav Med. 2023;13(2):122. 10.1093/tbm/ibac078] Transl Behav Med. 2022;12(8):878-883. 10.1093/tbm/ibac053

35. Ramanuj P, Ferenchik E, Docherty M, Spaeth-Rublee B, Pincus HA. Evolving models of integrated behavioral health and primary care. Curr Psychiatry Rep. 2019;21(1):4. 10.1007/s11920-019-0985-4

36. Miller SC, Frogner BK, Saganic LM, Cole AM, Rosenblatt R. Affordable Care Act impact on community health center staffing and enrollment: a cross-sectional study. J Ambul Care Manage. 2016;39(4):299–307. 10.1097/jac.0000000000000122

37. Wong SYS, Zhang D, Sit RWS, Yip BHK, Chung RYN, Wong CKM, et al. Impact of COVID-19 on loneliness, mental health, and health service utilisation: a prospective cohort study of older adults with multimorbidity in primary care. Br J Gen Pract. 2020;70(700):e817–e824. 10.3399/bjgp20x713021

38. Britz JB, Huffstetler AN, Henry TL, Ragunanthan B, Britton E, Doshi N, et al. Primary care: a critical stopgap of mental health services during the COVID-19 pandemic. J Am Board Fam Med. 2022;35(5):891–896. 10.3122/jabfm.2022.05.210523

